# Sedatives reduce human gastric slow-wave amplitude, assessed by a multicenter study of endoscopic gastric mapping

**DOI:** 10.64898/2026.10.15.26365429

**Authors:** Peter Tremain, Jarrah M. Dowrick, Chih-Hsiang Alexander Chan, David Rowbotham, Gary Lim, Gregory O’Grady, Leo K. Cheng, James Mckeage, Timothy R. Angeli-Gordon

**Author notes:** **Corresponding Author:** Dr. Timothy R. Angeli-Gordon, Auckland Bioengineering Institute, University of Auckland, Private Bag 92019, Auckland, New Zealand.

## Abstract

**Background:** Gastrointestinal (GI) motility disorders are prevalent, impose a significant burden, and are associated with disrupted spatial propagation of bioelectrical gastric slow waves. High-resolution endoscopic gastric mapping (EGM) is an emerging minimally invasive technique for imaging gastric electrical activity; however, EGM is performed alongside standard gastroscopy and requires procedural sedation, which may interact with gastric electrical activity, distorting clinical insights. This study explored the influence of procedural sedation on EGM-recorded slow-wave activity.

**Methods:** EGM was performed for ∼7 min in 32 patients undergoing upper GI endoscopy at two New Zealand hospitals. Endoscopists administered procedural sedation (fentanyl and midazolam) according to best clinical practice. The relationship between slow-wave amplitude and patient characteristics was then investigated using a variety of statistical techniques.

**Key Results:** Gastric slow waves were detected in 30 of 32 patients who underwent EGM. Symptom scores and demographic measures were comparable between sites. Fentanyl (A, 93.4 ± 13.7 μg vs B, 137.5 ± 33.7 μg, *p*≤0.001) and midazolam (A, 5.0 ± 0.9 mg vs B, 7.4 ± 1.6 mg, *p*≤0.001) doses were higher at Hospital B than Hospital A. Correspondingly, slow-wave amplitudes were lower at Hospital B than Hospital A (A, 2.33 ± 1.61 mV vs B, 0.86 ± 1.01 mV, *p*=0.004). All other slow-wave metrics were consistent between hospitals. Combined sedative dose was negatively correlated with slow-wave amplitude (*p*=0.037).

**Conclusions & Inferences:** Higher procedural sedative doses are associated with lower-amplitude gastric slow-waves. Procedural sedation dose and opiate administration should be carefully considered when interpreting GI electrophysiology measurements.

## 1. Introduction

Gastrointestinal (GI) motility disorders, such as gastroparesis (GP) and functional dyspepsia (FD), affect approximately 40% of the global population.^1^ These disorders can inflict debilitating symptoms, such as nausea, vomiting, abdominal pain, and heartburn, which severely impact quality of life.^2–4^ The financial burden these disorders impose on society is significant. These symptoms result in a higher unemployment rate among patients with GI motility disorders,^2,5^ and in the United States, the cost to treat GP and FD alone is almost USD$20 billion.^6,7^

Despite their prevalence, GI motility disorders remain poorly understood, with clinicians relying on patient-reported symptom burden and labile biomarkers, such as gastric emptying, for diagnosis.^8–10^ Consequently, diagnostic pathways are often prolonged and indeterminate, with an average time to GP diagnosis of 5 years.^11^ A form of bioelectrical activity in the gut, called slow waves, could serve as a more accurate biomarker of GI dysfunction.^12–14^ In health, slow waves propagate distally from the upper corpus toward the pylorus in a regular, coordinated pattern at a frequency of 2-4 cycles per minute (cpm).^15^ Conversely, motility disorders are associated with abnormal retrograde slow-wave propagation,^13,14,16^ along with amplitudes, frequencies, or velocities outside normal ranges.^17^ As a result, new electrophysiological tools are being developed to improve patient care.

Among these emerging GI electrophysiology tools, body-surface gastric mapping (BSGM) has reached the clinic and is proving a valuable diagnostic aid.^12,18,19^ However, the physical separation between the stomach and BSGM electrodes limits the spatial fidelity of these recordings.^20^ This limitation is particularly relevant for nascent targeted therapeutics, such as gastric ablation, which require accurate localization of abnormal electrical initiation sites.^21–23^ Although organ-surface electrical mapping provides enhanced spatial insights, conventional serosal methods have required surgical access under general anesthesia, limiting their clinical feasibility.^14,24^ Endoscopic gastric mapping (EGM) addresses this challenge by enabling high-resolution measurements of slow-wave propagation from the mucosal surface under conscious sedation.^25^

Organ-surface recordings obtained with EGM exhibit greater slow-wave amplitude than prior slow-wave recordings obtained with invasive methods.^25^ It was theorized that the greater slow-wave amplitude was partly due to the lower sedative dose administered during endoscopic recordings. This conclusion is supported by preclinical findings,^26–28^ but no human data have definitively established a relationship between sedative dose and slow-wave amplitude.

In this study, we investigated the relationship between procedural sedative dose and slow-wave amplitude from EGM performed at two different hospitals by two different endoscopists. Understanding the influence of sedation on slow-wave activity, including constituent opiates, has direct implications for the diagnosis of GI motility disorders and is essential for accurate interpretation of EGM electrical measurements.

## 2. Methods

This study utilized a recording device and method previously described by Tremain et al. to obtain slow-wave data from human participants undergoing upper-GI endoscopy.^25^ The device construction and recording procedure are summarized in brief below. Study design and data analysis methods are then described in full.

### 2.1 Recording Procedure Summary

The HR EGM device consisted of a modified 64-electrode basket-type electrical recording catheter (Constellation cardiac mapping catheter; Boston Scientific, Boston, MA), and a non-latex, anorectal manometric catheter inflation balloon (Mui Scientific, Mississauga, ON, Canada).

Sedation was administered by the endoscopist prior to delivery of the EGM device to ensure adherence to best practices in patient care. The EGM device was then delivered endoscopically to the gastric antrum, alongside a gastroscope (Fig. 1). Appropriate positioning in the gastric antrum was confirmed by visual inspection with the gastroscope. The EGM balloon was then inflated with 110 mL of air, using a 60-mL syringe, and the stomach was suctioned, using the gastroscope, to improve contact between the recording electrodes and the gastric mucosa. The EGM device was then left to record for ∼7 min before being deflated and removed.

**Figure 1:**
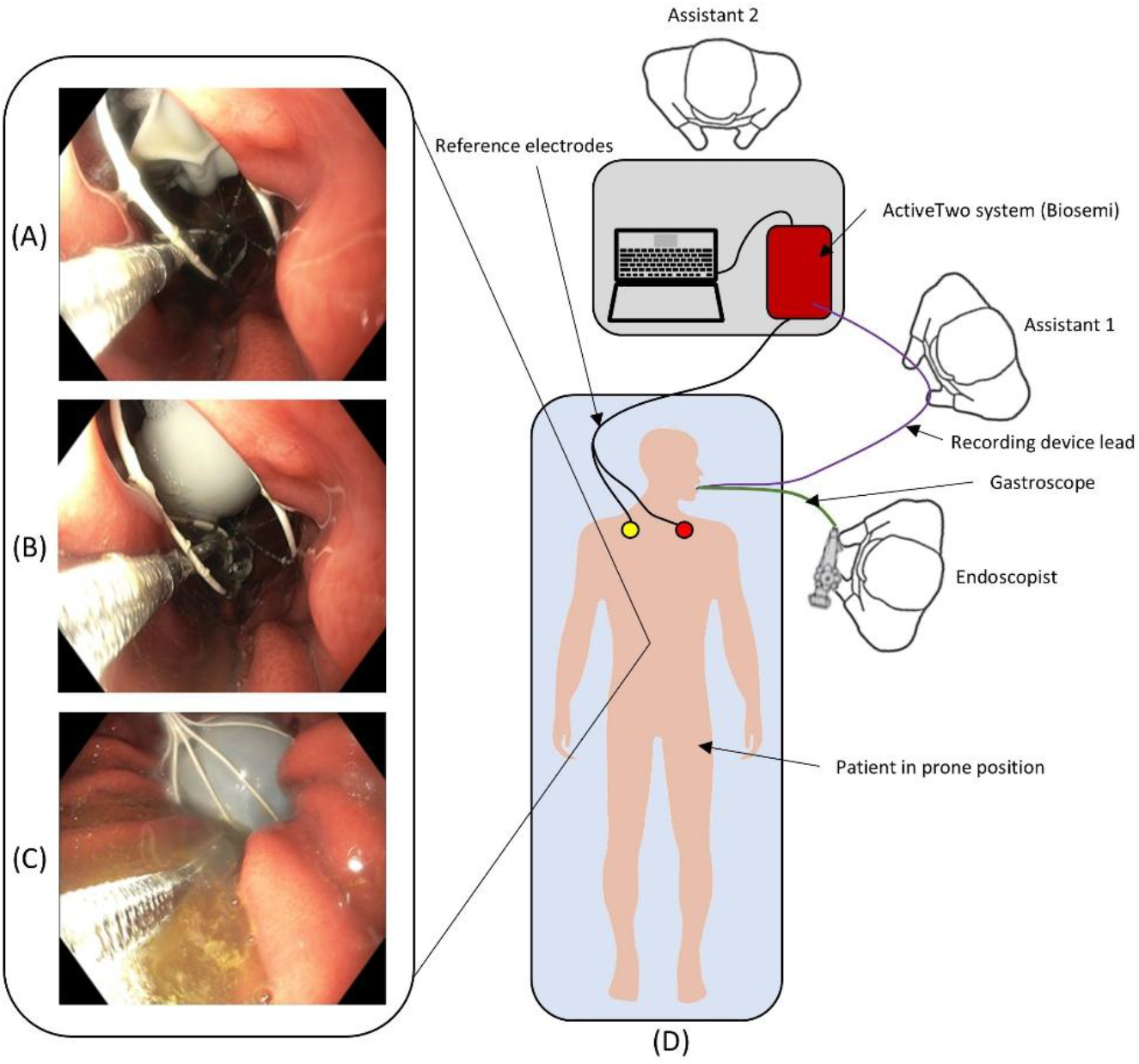
Setup of endoscopy suite and representative images of the device deployed in a patient (A-C). The patient was positioned prone with grounding leads adhered to the shoulder. (A) Balloon is in a fully deflated state, minimizing overall device size to make delivery of the device down the esophagus easier. (B) Balloon starting to inflate. (C) Balloon fully inflated in the gastric antrum with stomach suctioned down onto the device, ready to begin recording. (D) Endoscopy suite layout during procedure. Equipment was laid down during the recording period to minimize potential artifacts from equipment handling. Reproduced from Tremain et al. (2024).^25^

### 2.2 Clinical Study Design – Ethics Screening and Patient Selection

Ethical approval was granted by the Southern Health and Disability Ethics Committee (approval no. 17/STH/95). All participants provided informed consent, and the EGM data were deidentified via pseudonymization. Patients aged 18–75 years old, scheduled for upper GI endoscopy at two hospitals (Hospital A and Hospital B), were screened for inclusion in the study. The exclusion criteria focused on factors that could contribute to abnormal electrical propagation beyond functional causes, and conditions that put the patient at unnecessary risk during the procedure, including cirrhosis, pregnancy, previous gastric surgery, or use of medications known to alter gastric motility, along with patients using a pacemaker. Individuals who preferred not to have sedation for their scheduled endoscopy were also excluded to minimize the risk of excessive patient discomfort. Before the procedure, patient symptoms were evaluated using a validated questionnaire, the Patient Assessment of Upper Gastrointestinal Symptom Severity Index (PAGI-SYM).^29^ Classification into various disease groups was carried out after diagnosis and confirmed by gastroscopy immediately following the recording procedure. A review of the patient’s health records was also carried out to support disease classification.

### 2.3 Signal Acquisition, Processing, and Analysis

Signals were recorded simultaneously from all 64 electrodes, at 512 Hz, using an ActiveTwo™ system (BioSemi, Amsterdam, the Netherlands), modified for passive recordings. Signal processing and data analysis, including calculation of slow-wave frequency, amplitude, and velocity, were performed in custom software (Bezoar)^30,31^ developed specifically for endoscopic gastric electrical mapping data, running in MATLAB version R2021b (MathWorks, Natick, MA). Signals were downsampled to 32 Hz and then filtered with a Gaussian moving median filter for baseline removal (20-s window). In addition to downsampling and Gaussian filtering, discrete wavelet transforms, and SG filters (1.7 s window, polynomial order 9) were applied to remove high-frequency noise and aid in slow-wave identification.^32,33^ Slow-wave activation times (AT) were manually marked based on qualitative assessment of signal morphology and the temporal relationship with the signal deflections of other channels, which is consistent with marking criteria for previously validated serosal slow-wave mapping studies.^32,34^ Slow-wave velocity was evaluated in Bezoar using a thin-plate spline-directed local fitting method. Only recordings with a maximum recorded slow-wave coverage of at least 19 channels (30%) were used for velocity calculations, to avoid instability from excessive interpolation. Slow-wave amplitude was calculated as the difference between the maximum and minimum signal voltage within a 3 s time window around a marked slow-wave activation time (AT) (1.5 s either side of the marked point). Frequency was defined as the median time between marked slow-wave ATs per 60 s.

The signal-to-noise ratio (SNR) of recordings was calculated using a previously validated method that segments the filtered data into signal and noise components.^35^ Segments associated with signal data were defined as all points within a 3-s time window around a marked slow-wave AT (1.5 s either side of the marked point). All data points outside the 3-s signal windows were classified as noise. Equation 1 was then used to compute SNR.

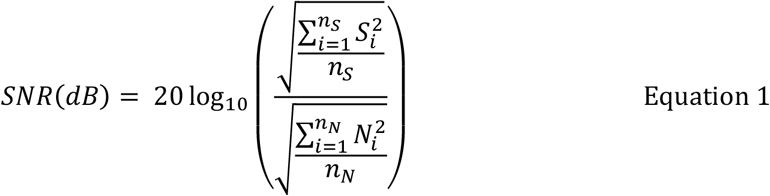

where *S_i_* is the signal data points, *n_S_* is the number of points included in the signal windows, *N_i_* is the noise data points, and *n_N_* is the number of points included in the noise windows of the recordings.

### 2.4 Statistical Analysis

When computing the amplitude, velocity, and frequency of each wavefront in a recording, the median was used to avoid the influence of physiological or mathematical outliers in the analysis. After median values for each quantitative measure were obtained for each wavefront, the mean and standard deviation (SD) were calculated for all wavefronts in a particular patient recording. When evaluating summative quantitative measures for the overall patient cohort or a particular subset of patients, the mean of means was calculated, along with an SD of the individual patient means.

Statistical analysis was performed in MATLAB version R2021b and Microsoft Excel version 2208. The normality of data distribution was assessed using a Shapiro-Wilk test. To analyze the statistical significance of relationships in discrete data types between the two hospitals (e.g., amplitude, frequency, age, etc.), an unpaired, two-tailed Welch’s t test was used if the distribution was normal. If the distribution was non-normal, then a Wilcoxon rank-sum test was used.

A secondary analysis was also carried out to investigate the influence of sedation and patient variation on slow-wave amplitude. This secondary analysis was conducted using R 4.5.2 with the *lme4* and *lmerTest* packages. To assess potential clustering by recruitment site, an initial linear mixed-effects model with hospital as a random intercept was fitted, and the random-effect term was evaluated using likelihood-ratio testing (LRT). The hospital random effect was then dropped, and various linear models were compared (detailed below) using the Bayesian Information Criterion (BIC), with a lower BIC indicating a more parsimonious model. The normality of the residuals from the most parsimonious model was assessed using the Shapiro-Wilk test and a visual inspection of the Q-Q plot. The covariate combinations used for the linear models were:

1) Sedative score + sex + age + overall PAGI-SYM score + body mass index (BMI)

2) Sedative score + sex + age + overall PAGI-SYM score

3) Sedative score + sex + age

4) Sedative score + sex

5) Sedative score

A combined sedative score was used because fentanyl and midazolam were administered together, and Pearson correlation analysis indicated that their dosages were highly correlated (⍴ > 0.7). Sedative score was defined as the sum of z-score-scaled fentanyl and midazolam doses.

Finally, analysis was conducted to elucidate any potential correlations between specific sub-symptom prevalence and slow-wave amplitude. The sub-symptoms considered were nausea, early satiety, bloating, upper abdominal pain, lower abdominal pain, and heartburn. Potential correlations were investigated using univariate linear regression and Spearman’s rank correlation, with *p*-values adjusted using Benjamini-Hochberg false discovery rate (FDR). Statistical significance for all tests was defined as *p* < 0.05.

## 3. Results

### 3.1 Summary of Recruited Patients

A total of 35 participants were recruited for the study. Three recruited patients withdrew after consenting but prior to recording, leaving 32 patients in whom recordings were performed (19 female, 13 male). A total of 19 recordings were carried out in Hospital A and 13 carried out in Hospital B. The mean age of patients in the study was 43.3 ± 14.4 years (range, 18-75 years), with no significant difference between study sites (A: 45.0 ± 13.1 years vs B: 40.9 ± 15.8 years, *p* = 0.45). The mean BMI was 27.6 ± 7.5 kg/m^2^ (range, 15.0–51.0 kg/m^2^), with no significant difference between study sites (A: 25.8 ± 4.8 kg/m^2^ vs B: 29.46 ± 9.11 kg/m^2^, *p* = 0.23). PAGI-SYM questionnaires assessing symptom severity before endoscopy returned a mean total score of 1.7 ± 0.9 on a scale of 0 (no symptoms) to 5 (very severe symptoms). No significant difference in PAGI-SYM scores was observed between study sites (A: 1.7 ± 0.9 vs B: 1.7 ± 0.8, *p* = 0.92) (Table 1).

**Table 1:** PAGI-SYM patient symptom scores were consistent between cohorts. Scores are reported for each study site and the overall study cohort. The p-values reported are for the relationship between the Hospital A and Hospital B study sites for the symptom group presented in each column.

| Symptom | Cohort |  |  | p-value |
| --- | --- | --- | --- | --- |
|  | Hospital A | Hospital B | All Patients |  |
| Heartburn | $1.4 \pm 0.9$ | $1.3 \pm 1.2$ | $1.4 \pm 1.0$ | 0.49 |
| Nausea | $0.7 \pm 0.7$ | $0.9 \pm 1.2$ | $0.8 \pm 1.0$ | 0.50 |
| Early Satiety | $1.8 \pm 1.1$ | $2.1 \pm 1.4$ | $1.9 \pm 1.2$ | 0.51 |
| Bloating | $2.5 \pm 1.8$ | $2.1 \pm 1.4$ | $2.4 \pm 1.7$ | 0.74 |
| Upper Abdominal Pain | $2.2 \pm 1.4$ | $2.4 \pm 1.4$ | $2.3 \pm 1.4$ | 0.35 |
| Lower Abdominal Pain | $1.7 \pm 1.3$ | $1.2 \pm 1.4$ | $1.5 \pm 1.4$ | 0.89 |
| Overall | $1.7 \pm 0.9$ | $1.7 \pm 0.8$ | $1.7 \pm 0.9$ | 0.92 |

The most common indications from endoscopy were forms of dyspepsia (*n* = 16 patients, A: 9 patients vs B: 7 patients). A breakdown of individual patient PAGI-SYM symptom scores is available in supplementary table S1, and a breakdown of individual patient indications following endoscopy is available in supplementary table S2.

### 3.2 Medications administered

Medications administered to patients during the study procedure included Xylocaine spray to numb the throat and intravenous conscious sedation (fentanyl 75–200 μg and midazolam 4–10 mg). The mean quantity of fentanyl administered during the procedure was 112.1 ± 32.6 μg, but this differed significantly between study sites, with more fentanyl being administered to patients in Hospital B (A: 93.4 ± 13.7 μg vs B: 137.5 ± 33.7 μg, *p* ≤0.001). The mean midazolam dose administered was 6.1 ± 1.7 mg, but again, higher doses were given to patients in Hospital B than in Hospital A (A: 5.0 ± 0.9 mg vs B: 7.4 ± 1.6 mg, p ≤0.001). A full list of medications administered to each patient is available in supplementary table S2.

### 3.3 Slow-Wave Recording Metrics, Traces, and Maps

Slow waves were successfully detected in 30 of the 32 recordings obtained. Of these 30 successful recordings, 17 were from Hospital A and the remaining 13 were from Hospital B. Sample recordings and propagation maps are shown in Figure 2.

**Figure 2:**
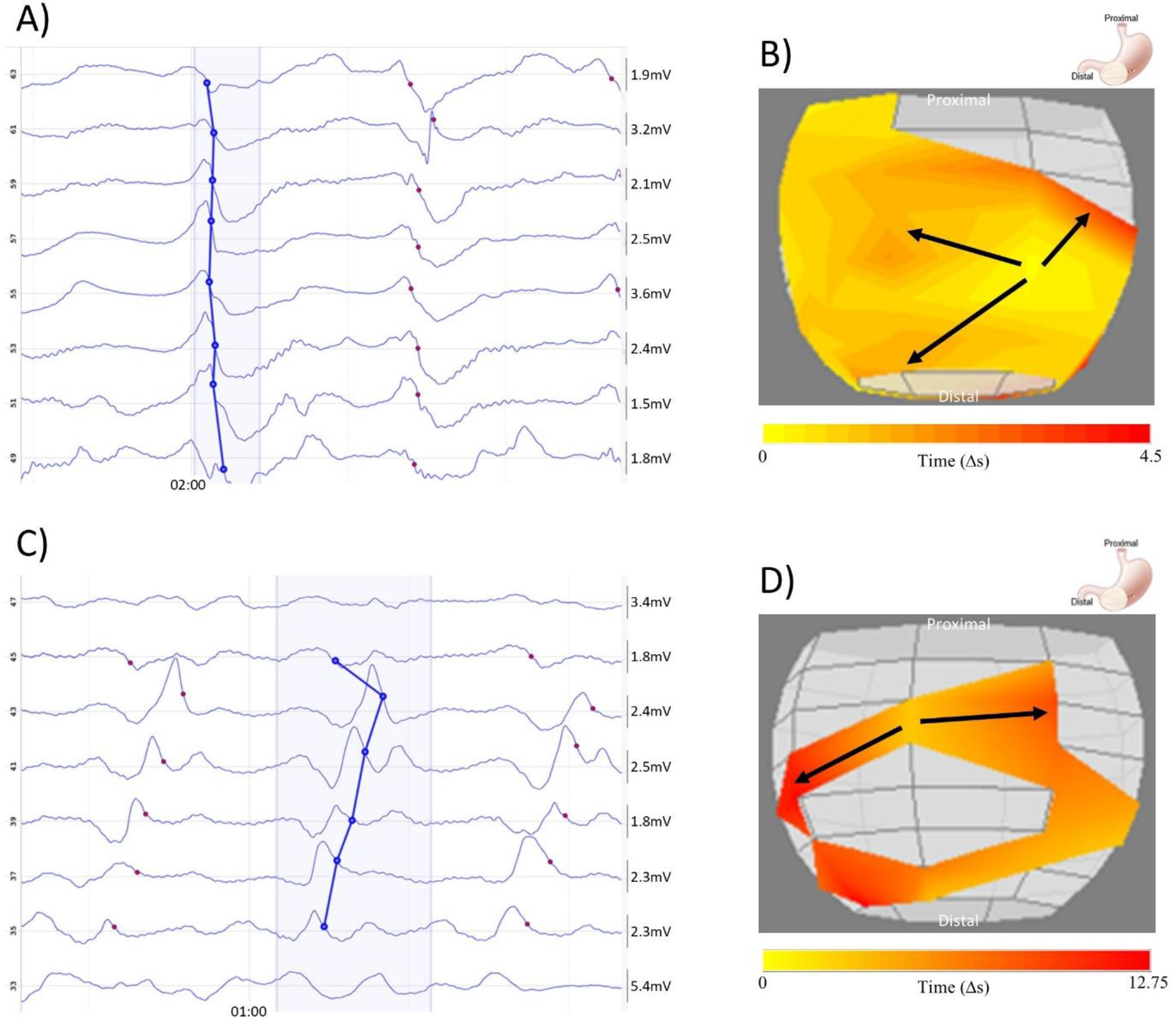
Human gastric electrical activity captured by the endoscopic mapping device. A & C) Sample signal traces from patient recordings. Slow-wave activations are marked by red dots, with the individual wave groups bounded within the blue shaded areas. B & D) Sample propagation maps of slow-wave activity, color bands represent 0.25 s intervals, black arrows indicate general slow-wave propagation direction.

In total, 12 recordings met the minimum coverage threshold for velocity calculations, 7 in Hospital A and 5 in Hospital B. The remaining slow-wave measures were calculated for all 30 successful recordings. With the exception of amplitude, which was greater at Hospital A (Fig. 3), slow-wave measures were consistent between study sites (Table 2).

**Figure 3:**
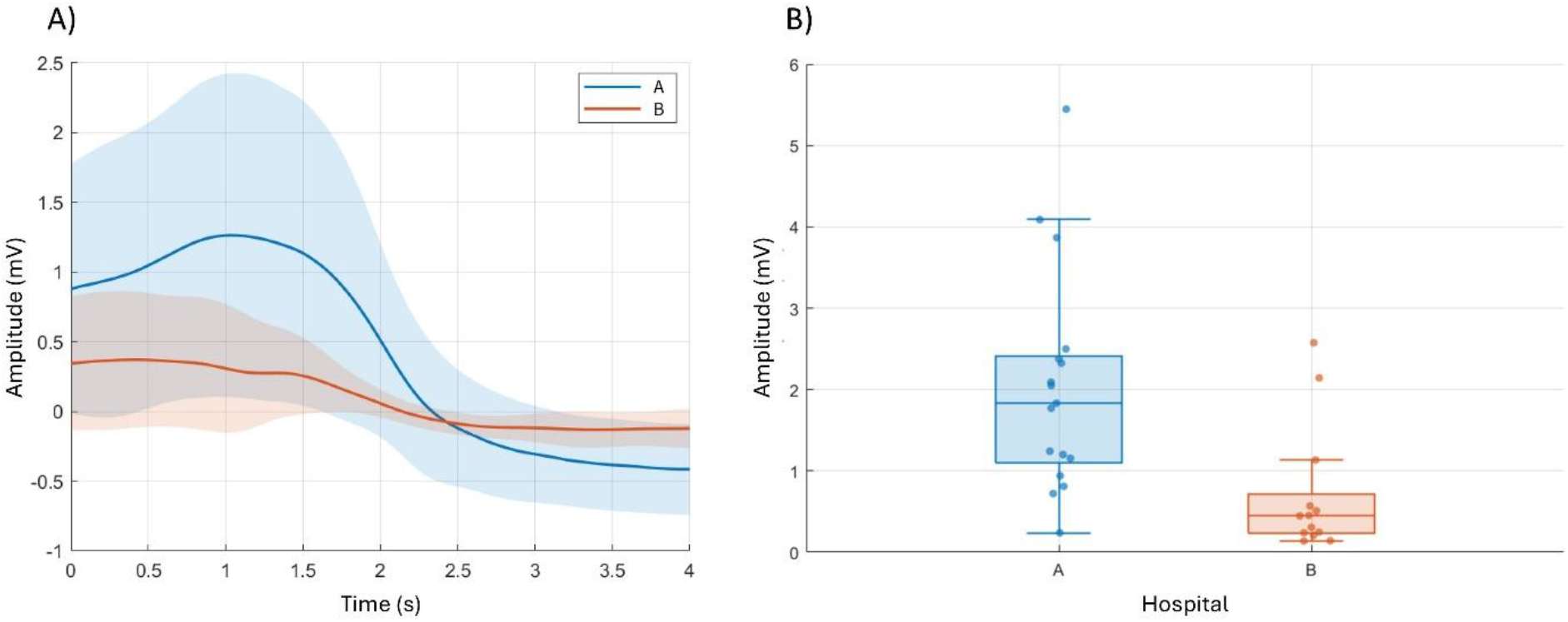
Slow-wave amplitude was lower at Hospital B than at Hospital A. A) Cohort-wise mean slow-wave activation signals (dark lines). Shaded regions represent the standard deviation of the mean signal. B) Box plot comparing mean slow-wave amplitudes from each hospital cohort. Hospital A is plotted in blue, and Hospital B is plotted in orange for both panels.

**Table 2:** Summary of slow-wave quantitative metrics. Metrics are reported for each study site and the overall study cohort. The p-values compare the Hospital A and Hospital B study sites for the metric presented in each row.

| Metric | Cohort |  |  | p-value |
| --- | --- | --- | --- | --- |
|  | Hospital A | Hospital B | All Patients |  |
| Coverage (maximum %) | 26.93 ± 21.16 | 18.03 ± 12.61 | 23.07 ± 18.50 | 0.23 |
| SNR (dB) | -0.92 ± 4.80 | -9.24 ± 18.27 | -4.41 ± 12.80 | 0.13 |
| Frequency (cpm) | 2.90 ± 0.42 | 2.69 ± 0.79 | 2.81 ± 0.60 | 0.42 |
| Amplitude (mV) | 2.33 ± 1.61 | 0.86 ± 1.01 | 1.71 ± 1.56 | 0.004 |
| Velocity (mm/s) | 5.09 ± 1.80 | 4.69 ± 2.84 | 4.92 ± 2.18 | 0.79 |

Analysis using the linear mixed-effects model indicated that the hospital random intercept was not supported (LRT = 0.498, *p* = 0.480); as such, the linear models without the hospital random effect are relied upon for further conclusions. The linear model with the lowest BIC, and consequently the most parsimonious model, was the univariate analysis of sedative score alone (BIC = 82.76). The BIC for alternate models ranged from 85.53 to 93.03 (supplementary table S3). This model indicated that slow-wave amplitude had a significant, negative association with sedative score (β = -0.261, *p* = 0.037). Shapiro-Wilk testing of the residuals indicated deviation from normality (W = 0.920, *p* = 0.027), and inspection of the Q-Q plot confirmed this was driven by a pronounced upward curvature in the upper tail (Fig. 4). While this suggests the presence of right-skewness in the residual distribution, given the sample size, the model was deemed sufficiently robust under the Central Limit Theorem.

**Figure 4:**
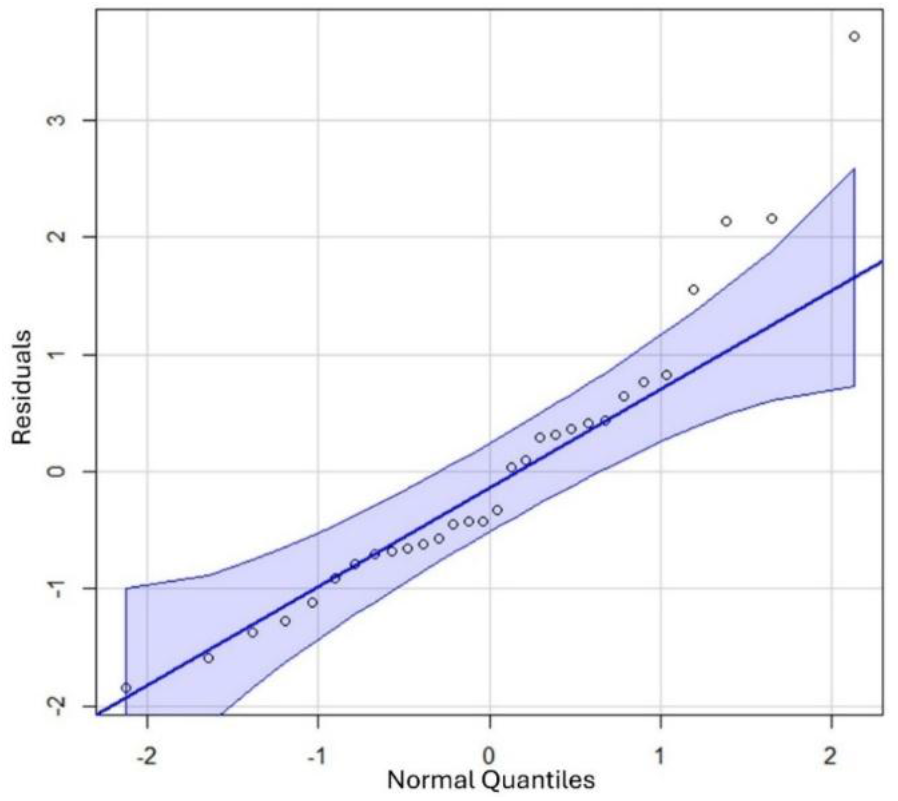
Q-Q plot of residuals from the linear model with the lowest BIC. Points show observed residual quantiles against theoretical normal quantiles. The shaded region indicates the 95% confidence interval.

Univariate sub-symptom analysis indicated no significant associations between any sub-symptoms and slow-wave amplitude (Table 3).

**Table 3:** Summary of linear regression modelling and Spearman correlation analysis of individual symptom influence on slow-wave amplitude.

| Symptom | Linear Regression |  |  |  | Spearman Correlation |  |  |
| --- | --- | --- | --- | --- | --- | --- | --- |
| | $\beta$ | SE | $p$ | $p(\text{FDR})$ | $\rho$ | $p$ | $p(\text{FDR})$ |
| Nausea | -0.366 | 0.236 | 0.132 | 0.395 | -0.256 | 0.173 | 0.884 |
| Early satiety | -0.217 | 0.204 | 0.298 | 0.470 | -0.094 | 0.621 | 0.884 |
| Bloating | -0.033 | 0.148 | 0.827 | 0.827 | 0.028 | 0.884 | 0.884 |
| Upper abdominal pain | -0.287 | 0.180 | 0.122 | 0.395 | -0.071 | 0.709 | 0.884 |
| Lower abdominal pain | -0.043 | 0.190 | 0.823 | 0.827 | -0.043 | 0.820 | 0.884 |
| Heartburn | -0.237 | 0.231 | 0.313 | 0.470 | -0.078 | 0.683 | 0.884 |

### 3.4 Lack of adverse events

Deployment of the EGM device during the recording procedure outlined in this study was achieved safely in all patients. No significant adverse events were experienced during the study.

## 4. Discussion

This multicenter clinical study demonstrates that procedural sedation dose impacts gastric slow-wave amplitude. The effect of decreased slow-wave amplitude was highly specific, with no other slow-wave metric influenced by procedural sedation, and no relationship was observed from exploratory analysis of slow-wave and symptom measures. This is also the first multicenter study of the EGM device, demonstrating the feasibility of this technology and technique for recording in a clinical setting.

The observed sedative-induced modulation of slow-wave activity in this study has immediate implications for existing gastric electrophysiological diagnostic aids, such as BSGM.^36^ Current BSGM workflows incorporate gastric slow-wave power analysis to identify patients with bioelectrical abnormalities.^36,37^ The recent Auckland Classification defines a low-amplitude phenotype;^38^ therefore, sedatives that reduce signal power independently of underlying pathophysiology may lead to false-positive results during testing, which is particularly relevant for the opiate component of the sedatives. BSGM is performed in the awake state, includes a meal as part of the test, and it is recommended that patients stop opiates before the recording. However, many patients struggle to stop the use of opiates, even for acute testing. The results of our present study reinforce the need to: i) continue to advise patients to cease the use of opiates prior to GI motility tests, and ii) where cessation is not possible, interpret such test results with caution. It remains to be seen whether the effects of opiates on slow-wave activity can be accounted for in the existing BSGM pipeline. In the meantime, the quantitative validation of sedative-induced modulation of slow-wave activity presented in this study can inform improved testing protocols and reduce the risk of misclassification.

Beyond BSGM, these data may also have implications for emerging direct-organ-contact electrophysiology technologies, which image the spatial propagation of slow waves to infer dysfunction.^39,40^ Accurate identification of the wavefronts required to generate these spatial activity maps requires a sufficient SNR. Hence, sedation-induced reductions in slow-wave amplitude may compromise the capacity to reliably resolve electrical events and, therefore, undermine the ability to detect gastric dysrhythmias. Emerging technologies like EGM may therefore need to be designed to reduce the need for sedation. For example, the EGM device and endoscope could be consolidated into a single, slimmer delivery package, reducing discomfort and, in turn, the need for sedation, as has been the case with new ultra-thin endoscopes.^41–43^ Emerging ingestible and implantable devices could provide a complementary approach for obtaining direct-organ-contact electrophysiological recordings without sedation.^26,44,45^ However, there are several limitations with these technologies that need to be addressed for clinical applications, including: (i) low spatial resolution, (ii) limited battery life, and/or (iii) data processing or transmission bandwidth.^46,47^ Should these limitations be addressed, a combination of EGM followed by lower-resolution ingestible or implantable monitoring may represent the future of direct-organ-contact electrophysiology technology in the GI tract.

Understanding the primary pharmacological driver of sedation-induced reduction in slow-wave amplitude may enable the design of sedation protocols that minimize this effect for future electrophysiological measurements. However, due to their collinearity, it was not possible to statistically isolate the contributions of midazolam and fentanyl. Both agents have plausible mechanistic links to reduced gastric electrical activity. As an opioid, fentanyl acts on μ-opioid receptors on enteric neurons, leading to a reduction in acetylcholine release, thereby suppressing GI motility.^48^ Opiates have also been associated with increased symptom severity in patients with GI disorders.^49,50^ Midazolam, meanwhile, potentiates inhibitory GABA-A signaling in the dorsal motor nucleus of the vagus, which projects to the stomach.^51^ This is slightly complicated by the fact that GABA-A signaling is excitatory in the enteric nervous system,^52^ but the net effect of midazolam appears to be to inhibit gastric motility.^53^ Additional research is ultimately required to determine the specific pharmacological driver of slow-wave modulation, building on the foundational observations in this present study.

Interestingly, symptom burden was not correlated with slow-wave metrics in our study. This finding remains exploratory due to the relatively modest sample sizes in this cohort, and the initial results contradict other studies that have shown that certain symptoms, such as nausea, are associated with changes in gastric electrical activity.^54–56^ Neither linear regression modeling nor Spearman correlation coefficients indicated significant relationships between slow-wave amplitude and any reported symptom category in our present study. However, these calculations should be considered alongside critical inspection of the absolute symptom burden. The cohort in this present study was not selected based on symptom burden, and the lack of correlation between symptoms and slow wave characteristics may likely be due to insufficient symptom presentation in this specific patient cohort. Further investigation with larger cohorts of higher symptom burden is warranted and could now be a focus of future applications of the EGM device.

This study was the first multicenter study of the EGM device, building on a smaller feasibility trial and reiterating the ability of this device to record clinically meaningful data.^25^ Limited electrode contact and coverage remain the primary challenges for this technology. To improve electrode contact and mapping coverage across the device, future design iterations could incorporate: (i) optimized electrode size, material, and geometry,^26,35^ (ii) specially designed runoff channels to reduce potential interference from gastric secretions on the mucosa, similar to those used in other GI devices for drug delivery,^57^ and (iii) anatomically-informed device shape to expand coverage across a larger region of the stomach and better match the gastric anatomy, similar to recent advancements in anatomically-specific devices for surgically-invasive recordings of the gastroduodenal junction.^58–60^ These potential solutions will be considered for future iterations of the EGM device.

In summary, minimally invasive, direct-organ, HR slow-wave recordings were successfully performed in 30 patients across two hospitals using an emerging EGM device. The results highlighted that increased levels of procedural sedation are associated with decreased slow-wave amplitude. Procedural sedation, and more broadly, opiates in general, should be avoided during GI electrophysiological recordings where possible, for example, with non-invasive BSGM. Future studies of GI electrophysiology in sedated patients and future device design in this field should account for the impact of procedural sedation to ensure that resultant analysis accurately reflects underlying pathophysiology.

## Data Availability

All data required to evaluate the conclusions in the presented study are included in the manuscript.

## Author contributions

Study concept and design: PT, JD, DR, GL, GOG, LKC, JM, TRA-G. Data collection: PT, JD, CAC, DR, GL, LKC, JM, TRA-G. Data analysis: PT, JD, CAC. Data interpretation: all authors. Drafting of manuscript: PT, TRA-G. Critical review of manuscript: all authors.

## Acknowledgments

We thank all of the participants, without whom this study would not have been possible. We also thank all research support staff and clinical staff involved in this study from the Surgical and Translational Research (STaR) Centre at the University of Auckland, Auckland City Hospital, and Christchurch Hospital.

## Supplementary Information

**Table S1:**
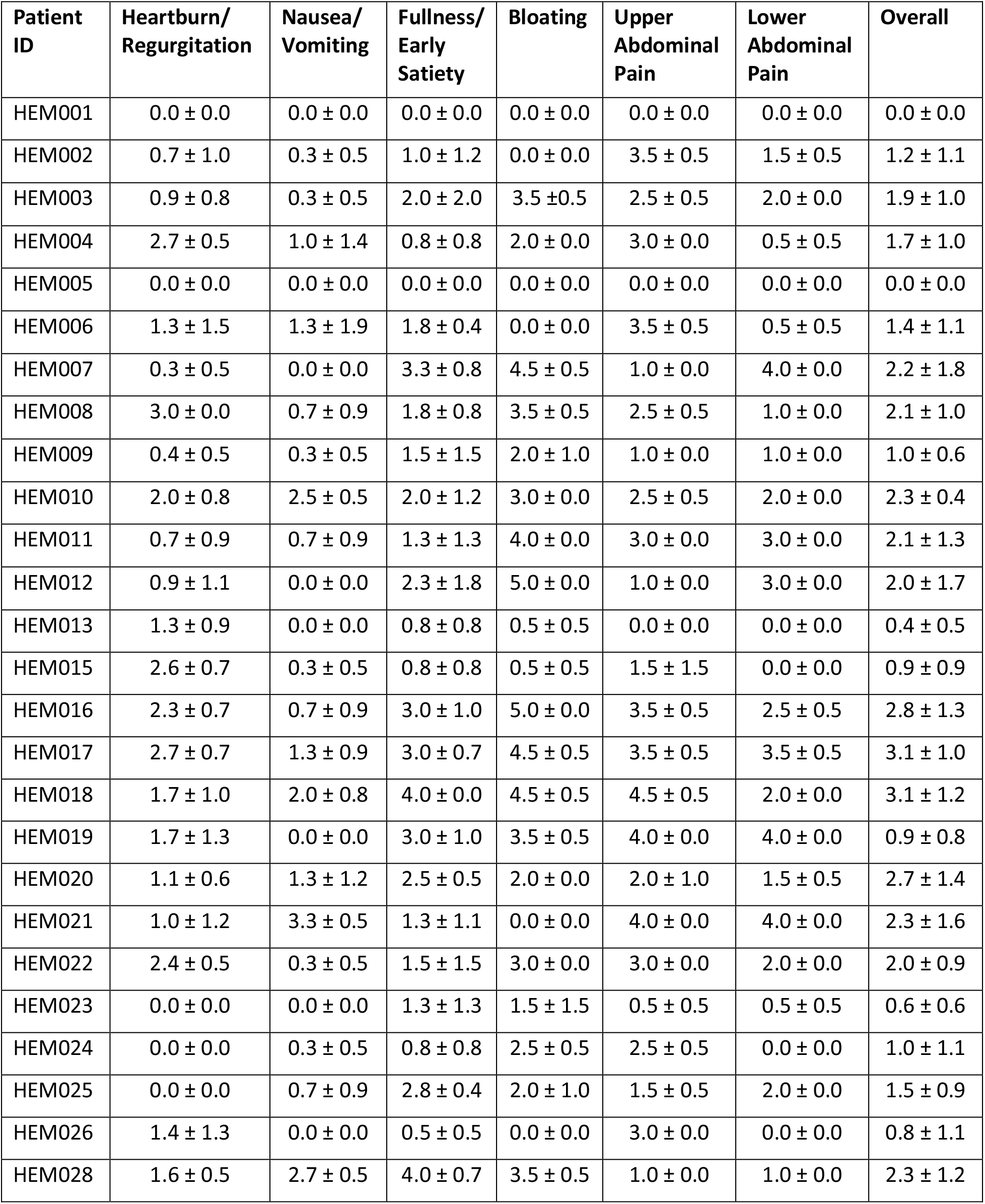
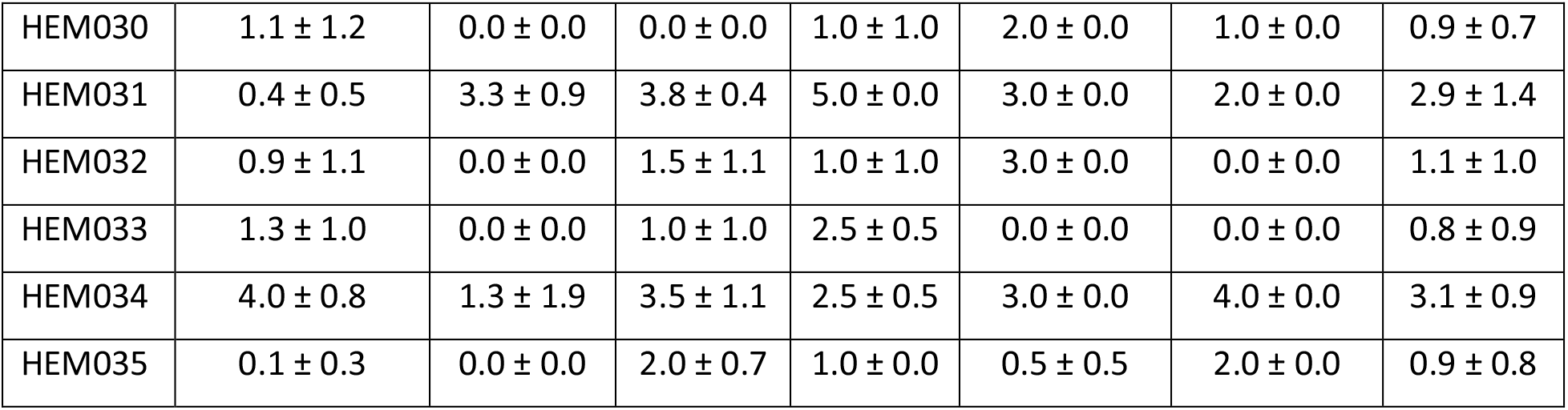
Summary of reported patient symptom scores as completed in pre-endoscopy PAGI-SYM questionnaires. HEM001 to HEM020 are Hospital A patients and HEM021 to HEM035 are Hospital B patients.

| Patient ID | Heartburn/<br>Regurgitation | Nausea/<br>Vomiting | Fullness/<br>Early Satiety | Bloating | Upper<br>Abdominal<br>Pain | Lower<br>Abdominal<br>Pain | Overall |
| --- | --- | --- | --- | --- | --- | --- | --- |
| HEM001 | 0.0 ± 0.0 | 0.0 ± 0.0 | 0.0 ± 0.0 | 0.0 ± 0.0 | 0.0 ± 0.0 | 0.0 ± 0.0 | 0.0 ± 0.0 |
| HEM002 | 0.7 ± 1.0 | 0.3 ± 0.5 | 1.0 ± 1.2 | 0.0 ± 0.0 | 3.5 ± 0.5 | 1.5 ± 0.5 | 1.2 ± 1.1 |
| HEM003 | 0.9 ± 0.8 | 0.3 ± 0.5 | 2.0 ± 2.0 | 3.5 ± 0.5 | 2.5 ± 0.5 | 2.0 ± 0.0 | 1.9 ± 1.0 |
| HEM004 | 2.7 ± 0.5 | 1.0 ± 1.4 | 0.8 ± 0.8 | 2.0 ± 0.0 | 3.0 ± 0.0 | 0.5 ± 0.5 | 1.7 ± 1.0 |
| HEM005 | 0.0 ± 0.0 | 0.0 ± 0.0 | 0.0 ± 0.0 | 0.0 ± 0.0 | 0.0 ± 0.0 | 0.0 ± 0.0 | 0.0 ± 0.0 |
| HEM006 | 1.3 ± 1.5 | 1.3 ± 1.9 | 1.8 ± 0.4 | 0.0 ± 0.0 | 3.5 ± 0.5 | 0.5 ± 0.5 | 1.4 ± 1.1 |
| HEM007 | 0.3 ± 0.5 | 0.0 ± 0.0 | 3.3 ± 0.8 | 4.5 ± 0.5 | 1.0 ± 0.0 | 4.0 ± 0.0 | 2.2 ± 1.8 |
| HEM008 | 3.0 ± 0.0 | 0.7 ± 0.9 | 1.8 ± 0.8 | 3.5 ± 0.5 | 2.5 ± 0.5 | 1.0 ± 0.0 | 2.1 ± 1.0 |
| HEM009 | 0.4 ± 0.5 | 0.3 ± 0.5 | 1.5 ± 1.5 | 2.0 ± 1.0 | 1.0 ± 0.0 | 1.0 ± 0.0 | 1.0 ± 0.6 |
| HEM010 | 2.0 ± 0.8 | 2.5 ± 0.5 | 2.0 ± 1.2 | 3.0 ± 0.0 | 2.5 ± 0.5 | 2.0 ± 0.0 | 2.3 ± 0.4 |
| HEM011 | 0.7 ± 0.9 | 0.7 ± 0.9 | 1.3 ± 1.3 | 4.0 ± 0.0 | 3.0 ± 0.0 | 3.0 ± 0.0 | 2.1 ± 1.3 |
| HEM012 | 0.9 ± 1.1 | 0.0 ± 0.0 | 2.3 ± 1.8 | 5.0 ± 0.0 | 1.0 ± 0.0 | 3.0 ± 0.0 | 2.0 ± 1.7 |
| HEM013 | 1.3 ± 0.9 | 0.0 ± 0.0 | 0.8 ± 0.8 | 0.5 ± 0.5 | 0.0 ± 0.0 | 0.0 ± 0.0 | 0.4 ± 0.5 |
| HEM015 | 2.6 ± 0.7 | 0.3 ± 0.5 | 0.8 ± 0.8 | 0.5 ± 0.5 | 1.5 ± 1.5 | 0.0 ± 0.0 | 0.9 ± 0.9 |
| HEM016 | 2.3 ± 0.7 | 0.7 ± 0.9 | 3.0 ± 1.0 | 5.0 ± 0.0 | 3.5 ± 0.5 | 2.5 ± 0.5 | 2.8 ± 1.3 |
| HEM017 | 2.7 ± 0.7 | 1.3 ± 0.9 | 3.0 ± 0.7 | 4.5 ± 0.5 | 3.5 ± 0.5 | 3.5 ± 0.5 | 3.1 ± 1.0 |
| HEM018 | 1.7 ± 1.0 | 2.0 ± 0.8 | 4.0 ± 0.0 | 4.5 ± 0.5 | 4.5 ± 0.5 | 2.0 ± 0.0 | 3.1 ± 1.2 |
| HEM019 | 1.7 ± 1.3 | 0.0 ± 0.0 | 3.0 ± 1.0 | 3.5 ± 0.5 | 4.0 ± 0.0 | 4.0 ± 0.0 | 0.9 ± 0.8 |
| HEM020 | 1.1 ± 0.6 | 1.3 ± 1.2 | 2.5 ± 0.5 | 2.0 ± 0.0 | 2.0 ± 1.0 | 1.5 ± 0.5 | 2.7 ± 1.4 |
| HEM021 | 1.0 ± 1.2 | 3.3 ± 0.5 | 1.3 ± 1.1 | 0.0 ± 0.0 | 4.0 ± 0.0 | 4.0 ± 0.0 | 2.3 ± 1.6 |
| HEM022 | 2.4 ± 0.5 | 0.3 ± 0.5 | 1.5 ± 1.5 | 3.0 ± 0.0 | 3.0 ± 0.0 | 2.0 ± 0.0 | 2.0 ± 0.9 |
| HEM023 | 0.0 ± 0.0 | 0.0 ± 0.0 | 1.3 ± 1.3 | 1.5 ± 1.5 | 0.5 ± 0.5 | 0.5 ± 0.5 | 0.6 ± 0.6 |
| HEM024 | 0.0 ± 0.0 | 0.3 ± 0.5 | 0.8 ± 0.8 | 2.5 ± 0.5 | 2.5 ± 0.5 | 0.0 ± 0.0 | 1.0 ± 1.1 |
| HEM025 | 0.0 ± 0.0 | 0.7 ± 0.9 | 2.8 ± 0.4 | 2.0 ± 1.0 | 1.5 ± 0.5 | 2.0 ± 0.0 | 1.5 ± 0.9 |
| HEM026 | 1.4 ± 1.3 | 0.0 ± 0.0 | 0.5 ± 0.5 | 0.0 ± 0.0 | 3.0 ± 0.0 | 0.0 ± 0.0 | 0.8 ± 1.1 |
| HEM028 | 1.6 ± 0.5 | 2.7 ± 0.5 | 4.0 ± 0.7 | 3.5 ± 0.5 | 1.0 ± 0.0 | 1.0 ± 0.0 | 2.3 ± 1.2 |
| HEM030 | 1.1 ± 1.2 | 0.0 ± 0.0 | 0.0 ± 0.0 | 1.0 ± 1.0 | 2.0 ± 0.0 | 1.0 ± 0.0 | 0.9 ± 0.7 |
| HEM031 | 0.4 ± 0.5 | 3.3 ± 0.9 | 3.8 ± 0.4 | 5.0 ± 0.0 | 3.0 ± 0.0 | 2.0 ± 0.0 | 2.9 ± 1.4 |
| HEM032 | 0.9 ± 1.1 | 0.0 ± 0.0 | 1.5 ± 1.1 | 1.0 ± 1.0 | 3.0 ± 0.0 | 0.0 ± 0.0 | 1.1 ± 1.0 |
| HEM033 | 1.3 ± 1.0 | 0.0 ± 0.0 | 1.0 ± 1.0 | 2.5 ± 0.5 | 0.0 ± 0.0 | 0.0 ± 0.0 | 0.8 ± 0.9 |
| HEM034 | 4.0 ± 0.8 | 1.3 ± 1.9 | 3.5 ± 1.1 | 2.5 ± 0.5 | 3.0 ± 0.0 | 4.0 ± 0.0 | 3.1 ± 0.9 |
| HEM035 | 0.1 ± 0.3 | 0.0 ± 0.0 | 2.0 ± 0.7 | 1.0 ± 0.0 | 0.5 ± 0.5 | 2.0 ± 0.0 | 0.9 ± 0.8 |

**Table S2:** Summary of medications used during the recording procedure, along with disease diagnosis or indication resulting from the completed endoscopy. HEM001 to HEM020 are Hospital A patients and HEM021 to HEM035 are Hospital B patients.

| Patient ID | Medications Used During Endoscopy | Diagnosis/Indication Post-Endoscopy |
| --- | --- | --- |
| HEM001 | Xylocaine spray<br>Fentanyl 100 mg<br>Midazolam 5 mg | Anemia, dyspepsia |
| HEM002 | Xylocaine spray<br>Fentanyl 75 mg<br>Midazolam 4 mg | Dyspepsia, suspected GERD, weight loss |
| HEM003 | Xylocaine spray<br>Fentanyl 100 mg<br>Midazolam 4 mg | Suspected GERD, globus sensation |
| HEM004 | Xylocaine spray<br>Fentanyl 100 mg<br>Midazolam 5 mg | Barrett's esophagus |
| HEM005 | Xylocaine spray<br>Fentanyl 100 mg<br>Midazolam 5 mg | Epigastric abdominal pain |
| HEM006 | Xylocaine spray<br>Fentanyl 100 mg<br>Midazolam 5 mg | Suspected GERD |
| HEM007 | Xylocaine spray<br>Fentanyl 75 mg<br>Midazolam 5 mg | Iron deficiency anemia |
| HEM008 | Xylocaine spray<br>Fentanyl 100 mg<br>Midazolam 5 mg | Suspected GERD |
| HEM009 | Xylocaine spray<br>Fentanyl 100 mg<br>Midazolam 4 mg | Dyspepsia |
| HEM010 | Xylocaine spray<br>Fentanyl 100 mg<br>Midazolam 5 mg | Epigastric abdominal pain |
| HEM011 | Xylocaine spray<br>Fentanyl 100 mg<br>Midazolam 5 mg | Epigastric abdominal pain |
| HEM012 | Xylocaine spray | Positive coeliac serologies |
|  | Fentanyl 100 mg<br>Midazolam 6 mg |  |
| HEM013 | Xylocaine spray<br>Fentanyl 100 mg<br>Midazolam 6 mg | GERD |
| HEM015 | Xylocaine spray<br>Fentanyl 100 mg<br>Midazolam 5mg | Hiatal hernia with reflux esophagitis |
| HEM016 | Xylocaine spray<br>Fentanyl 100 mg<br>Midazolam 7.5mg | Hiatal hernia with reflux esophagitis |
| HEM017 | Xylocaine spray<br>Fentanyl 100 mg<br>Midazolam 5mg | Functional Dyspepsia |
| HEM018 | Xylocaine spray<br>Fentanyl 100 mg<br>Midazolam 6mg | Suspected pernicious anemia |
| HEM019 | Xylocaine spray<br>Fentanyl 50 mg<br>Midazolam 4mg | Dyspepsia |
| HEM020 | Xylocaine spray<br>Fentanyl 75 mg<br>Midazolam 4mg | Dyspepsia |
| HEM021 | Fentanyl 100mg<br>Midazolam 6mg | Reflux esophagitis |
| HEM022 | Fentanyl 125mg<br>Midazolam 7mg | Dyspepsia, reflux |
| HEM023 | Fentanyl 100mg<br>Midazolam 6mg | Dyspepsia, reflux |
| HEM024 | Fentanyl 125mg<br>Midazolam 7mg | Hiatal hernia |
| HEM025 | Fentanyl 175mg<br>Midazolam 8mg | Abdominal pain, bloating |
| HEM026 | Fentanyl 150mg<br>Midazolam 7mg | Dyspepsia |
| HEM028 | Fentanyl 150mg<br>Midazolam 9mg | Nausea, abdominal pain |
| HEM030 | Fentanyl 125mg<br>Midazolam 9mg | GERD |
| HEM031 | Fentanyl 200mg<br>Midazolam 9mg | Nausea, vomiting |
| HEM032 | Fentanyl 150mg<br>Midazolam 7mg | Dyspepsia |
| HEM033 | Fentanyl 100mg<br>Midazolam 5mg | Hiatal hernia |
| HEM034 | Fentanyl 200mg<br>Midazolam 10mg | GERD |
| HEM035 | Fentanyl 100mg<br>Midazolam 5mg | Dyspepsia |

**Table S3:** Summary of BIC values for linear regression models analyzing the influence of sedation and patient variation on slow-wave amplitude.

| **Model Covariate Combinations** | **BIC** |
| --- | --- |
| Sedative score + sex + age + overall Pagi-SYM score + body mass index (BMI) | 93.03 |
| Sedative score + sex + age + overall Pagi-SYM score | 90.60 |
| Sedative score + sex + age | 87.43 |
| Sedative score + sex | 85.53 |
| Sedative score | 82.76 |

